# Investigating the biological and technical origins of unknown bases in the S region of the SARS-CoV-2 Delta variant genome sequences

**DOI:** 10.1101/2021.09.09.21262951

**Authors:** Loïc Borcard, Sonja Gempeler, Miguel A. Terrazos Miani, Christian Baumann, Carole Grädel, Ronald Dijkman, Franziska Suter-Riniker, Stephen L Leib, Pascal Bittel, Stefan Neuenschwander, Alban Ramette

## Abstract

We are reporting on the observation of a large, under-sequenced region of the S gene of the SARS-CoV2 Delta variant genomes, identified in sequences originating from various sequencing centres worldwide (e.g. USA, India, England, Switzerland, France, Germany). This poorly sequenced region was identified from the early phases of the Delta variant spread and the phenomenon is still ongoing. As many commonly-used protocols rely on amplicon-based sequencing procedures, we investigated the likely origin of the issue. We established its biological origin as resulting from mutations in the viral genomes at primer binding sites. We designed and evaluated new PCR primers to circumvent this issue in order to complement the ARTIC v3 set, and validated their performance for the sequencing of circulating Delta variants.

## Introduction

There is a worldwide effort to monitor the emergence and evolution of SARS-CoV-2 genotypes, due to the implications these new variants have for public health, society and scientific research. Such efforts have permitted the discovery of newly emerging variants, such as the Alpha, Beta, and Delta variants among others, and to follow their progression in the human population. In July 2020, the Delta variant, corresponding to the Pangolin lineage B.1.617.2, was first detected in India. Since then, this variant has been detected in the majority of countries and has rapidly become dominant in most countries worldwide [1-3]. The emergence of new variants often relies on the appearance of mutations within the S region of SARS-CoV2, the binding protein of SARS-CoV2 and the predominant antigen of the virus [4, 5]. The Delta variant (B.1.617.2) is a lineage circulating with mutations of biological significance S:P681R and S:L452R, and as of 12 August 2021, it was present in 115 countries.

Whole-genome sequencing (WGS) currently represents the methodology of choice for lineage classification and for providing fine genomic comparisons among viral isolates from local to global scales, allowing their surveillance in an unprecedented close-to-real-time manner. The rapid near-full-length sequencing of the virus genome has been enabled by the combination of, on the one hand, protocols that used amplicon tilling strategies, whereby allowing the fast and reliable production of complete genome sequences based on sets of carefully chosen amplicons to cover most of the 29-kb long genome, and on the other hand, the generalized use of Next Generation sequencing (NGS) technologies to massively sequence the resulting amplicons via short-read or long-read NGS technologies [6, 7]. Among the often-used approaches, the ARTIC protocol has received a lot of attention, as it was among the first available early in the pandemic (12 February 2020). The ARTIC network pipeline comprises a set of materials to assist sequencing by providing laboratory protocols and bioinformatic protocols. Each of the >100 pairs of proposed PCR primers in the protocol produces an insert of approximately 400 bases. The bioinformatic analysis of the data is performed by first, filtering the reads, trimming primer sequences, and normalising read coverage. Then, variant calling and consensus building are performed. The consensus sequences are then taxonomically classified using NextClade [8] and, more recently, the dynamic approach of Pangolin lineages [9].

The reliance on a PCR-based strategy to rapidly sequence viral genomes presents, however, a major drawback. Amplicon sequencing is prone to coverage dropouts that result in incomplete genome coverage, which leads to the reporting of unknown bases (represented as “N”) in the consensus sequences and sometimes prevents the correct lineage classification of viruses. There is an inherent danger when relying on genome specific primers as the template used to create these is bound to change with the genetic evolution of the virus. Thus, there is a constant effort to prevent the emergence of under-sequenced regions (USR) which are due to mismatching primers resulting from mutations of the virus sequences.

The aims of the study were: i) to report on the presence of a large USR in the S region of the Delta variant genome sequences, ii) to investigate the extent of the phenomenon among all the currently available Delta sequences, iii) to address its origin, as to whether it is biological (i.e. deletion) or technical (poorer amplification), and iv) to propose some solutions to sequence the missing region.

## Methods

### Samples

SARS-COV-2 RNA samples analysed in the study were obtained during the routine sequencing of SARS-CoV-2 viral genomes done at the Institute for Infectious Diseases (IFIK, Bern, Switzerland) for diagnostic or surveillance purposes. Ethics approval was granted by the Cantonal ethical commission for Research, Canton of Bern, Switzerland, on 17 February 2020 (GSI-KEK, BASEC-Nr Req-2020-00167) to sequence and genomically compare SARS-CoV-2 isolates starting from previously screened samples sent to the IFIK for viral diagnostics by treating physicians. Representative RNA samples were collected from 17 to 25 July 2021 from Lambda and Delta variants. All test results were anonymized prior to statistical analysis. Genome sequence information of all isolates is already available in the GISAID database (https://www.epicov.org/; [10]). Total nucleic acids were extracted using the STARMag 96 X4 Universal Cartridge Kit on a Seegene STARlet liquid handling platform (Seegene Inc., Seoul, South Korea). Briefly, 200 µl of original material was extracted in 100 µl elution buffer according to the manufacturer’s instructions (IVD). Nucleic acid eluates were immediately stored at -80°C after processing until further use. To investigate the situation in other countries, we downloaded a total of 436,820 sequences from GISAID on 12 August 2021 consisting of all Delta variants (B1.167.2, and sub-lineages AY.1; AY.2; AY.3; AY.3.1). The GISAID database offers the most exhaustive collection of curated sequences of SARS-CoV-2 originating from patient samples.

### Bioinformatic analyses

The ROI (Region of Interest, i.e. positions 21357-22246 of SARS-CoV-2 reference genome MN908947.3) was selected in the FASTA sequences by performing in silico PCR with *seqkit* (v0.16, [11]) using the ROI flanking primers. The forward primer binds between positions 21,357-21,386 (**Table 1**) and the reverse primer to positions 22,324-22,346 of the reference genome sequence. The proportion of Ns in the ROI sequences was calculated with the *alphabetfrequency* function of the *Biostrings* package [12]. Next the total frequency of Ns was computed using the *fx2tab* function of *seqkit*. Simultaneously, we analysed the presence of segments of N within the entire genome of SARS-CoV-2 by using the *locate* function of *seqkit* to detect all segments of N within the genome. All of this information was merged into one single dataset.

**Table 1.**
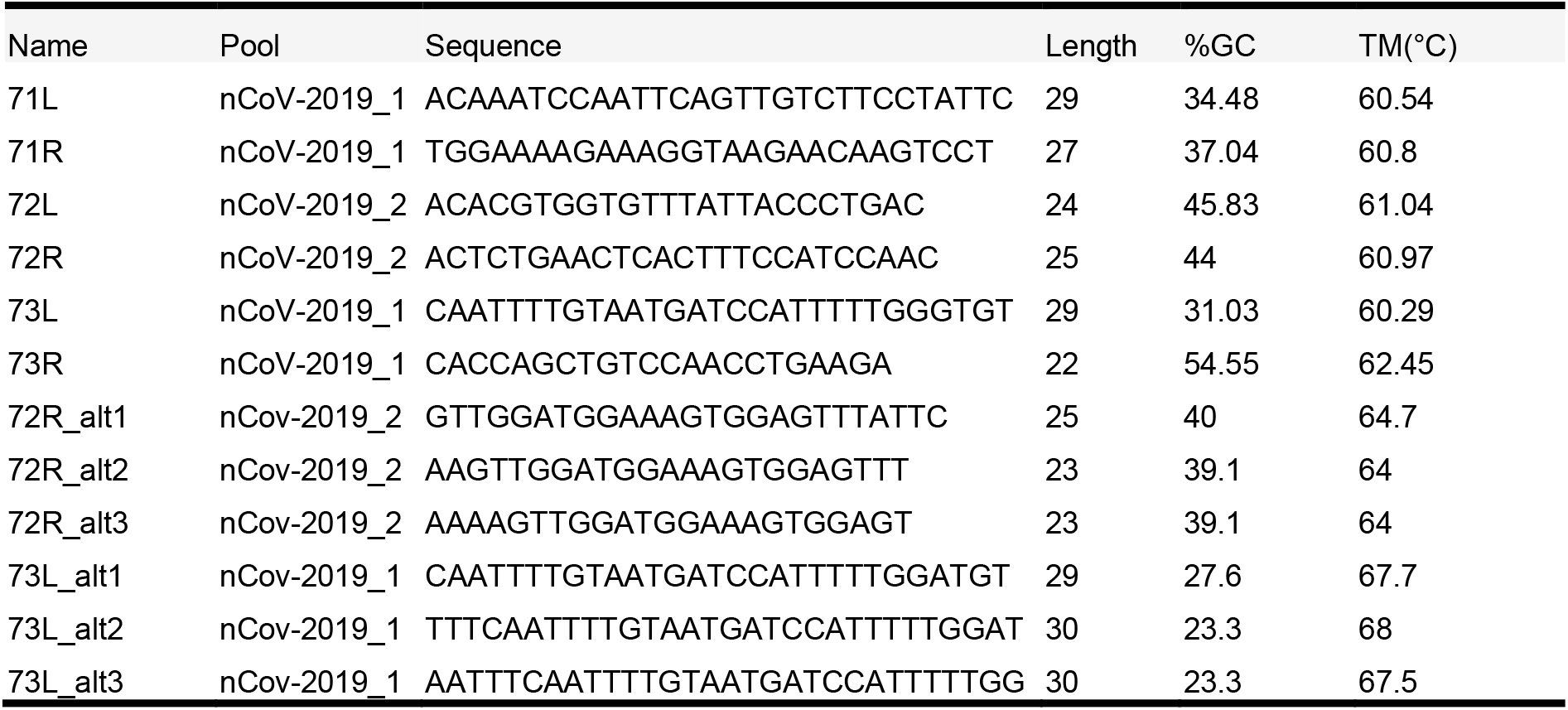
Original primers (ARTIC v3 protocol) and alternative primers binding to the ROI defined in this study.

### PCR and Sanger sequencing validation

The RT-PCR mix contained for each sample 16 µl RNA sample and 4 µl LunaScript RT Supermix (5x) (New England BioLabs, Ipswich, USA). The RT-PCR conditions were set to 2 min 25°C, 10 min 55°C, 1 min 95°C on a S1000 Thermal Cycler (Bio-Rad, Cressier, Switzerland). A PCR Mastermix with 12.5 µl Q5 Hot Start High-Fidelity 2x Master Mix (New England BioLabs), 1.85 µl of each 10 nM primers L and R (Microsynth AG, Balgach, Switzerland) and 3.8 µl nuclease-free water (New England BioLabs) was prepared per sample. A total of 20 µl of the PCR Mastermix was added to 5 µl of the RT sample. The PCR was set on a Bio-Rad S1000 Thermal Cycler with initial denaturation at 98°C (30s), followed by 35 amplification cycles of 98°C for 15 s and 65°C for 5 min. Gel electrophoresis was performed for 1 h at 120 volts with 2% agarose gels prepared in 1x TAE buffer with 2.5 µl RedSafe (20000x) (iNtRON Biotechnology, Burlington, USA). A total of 10 µl PCR reactions with DNA Loading buffer (ROTI Load DNA with Saccharose, Carl Roth GmbH) was loaded per well, alongside a Benchtop 100-bp DNA Ladder (Promega, Dübendorf, Switzerland). Gel images were acquired with the Fusio Fx (Vilber; Witec, Sursee, Switzerland) gel-documentation system. Purified PCR products were sent for Sanger sequencing at Microsynth. Forward and reverse strands were assembled into consensus sequences using SeqMan Pro (DNAStar, Madison, WI, USA) (Supplementary File 1.

### Nanopore sequencing

SARS-CoV-2 RNA genomes were reverse-transcribed, and amplified following the sequencing strategy of the ARTIC v3 protocol (https://artic.network/ncov-2019), which generates 400-bp amplicons that overlap by approximately 20 bp and covers the whole target genomes. Nanopore library preparation was performed with SQK-LSK109 (Oxford Nanopore Technologies, Oxford, UK) according to the ONT “PCR tiling of COVID-19 virus” (version: PTC_9096_v109_revE_06Feb2020, last update: 26/03/2020). Reagents, quality control and flow cell preparation were done as described previously [13, 14]. Sequencing was performed on GridION X5 (Oxford Nanopore Technologies) with real-time basecalling enabled (ont-guppy-for-gridion v.4.2.3; fast basecalling mode). Bioinformatic analyses followed the ARTIC workflow (https://artic.network/ncov-2019/ncov2019-bioinformatics-sop.html) using version 1.1.3. Consensus sequences were generated using *medaka* (https://github.com/nanoporetech/medaka) and *bcftools* [15].

## Results

### Evidence of a large USR resulting from mutations in the S gene

During routine sequencing of SARS-CoV-2 genomes at the Institute for Infectious Diseases in May 2021, we initially observed that the region surrounding the deletion 69-70 (nt positions) in the S region of SARS-CoV-2 Delta variant genome sequences (**Fig. 1**) was systematically under-sequenced (**Fig. 2A**), as compared to non-Delta variant sequences (**Fig. 2B**). The consensus sequences displayed a large region containing mostly N (unidentified) bases from positions 21,357 to 22,346 of the genome sequence and the inspection of the ROI coverage indicated the presence of only few to no reads in this region. The cause of the USR in the S gene was examined by PCR amplifying the ROI using USR-flanking primers 71L and 73R (**Fig. 2C**). PCR results confirmed the absence of a major deletion in this region, as the expected ca. 990-nt long region was amplified for all Delta and Lambda samples tested. To test the hypothesis of primer mismatch, PCR amplicons were further Sanger sequenced over positions 21,357-22,346 using the same two primers (71L and 73R) (**Fig. 2D**), and we could identify two non-matching primers: First, primer 72R (22013-22038) displayed a truncated binding site due to a deletion between position 22029-22034. Second, a substitution (G21987A) was detected in the primer binding site of 73L, which binds between position 21,961 to 21,990. These two primer sequence mismatches were found in all five consensus sequences considered here, supporting the hypothesis that the low coverage of the ROI was due to compromised binding of certain primers.

**Figure 1.**
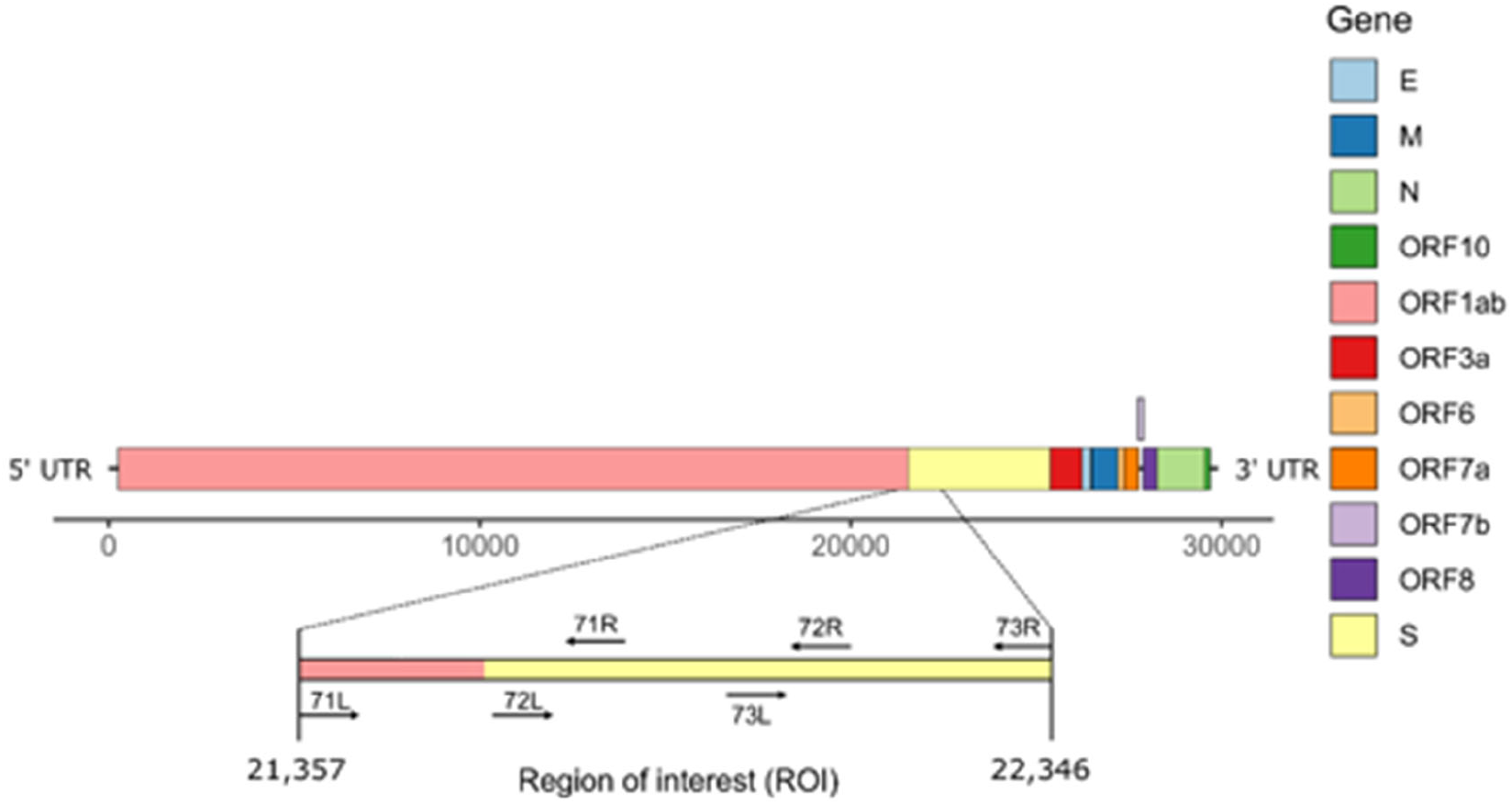
Schematic representation of SARS-CoV-2 genome structure. The region of interest (ROI) at positions 21,357-22,346 corresponds to the region between primers 71L and 73R (ARTIC primers v3).

**Figure 2.**
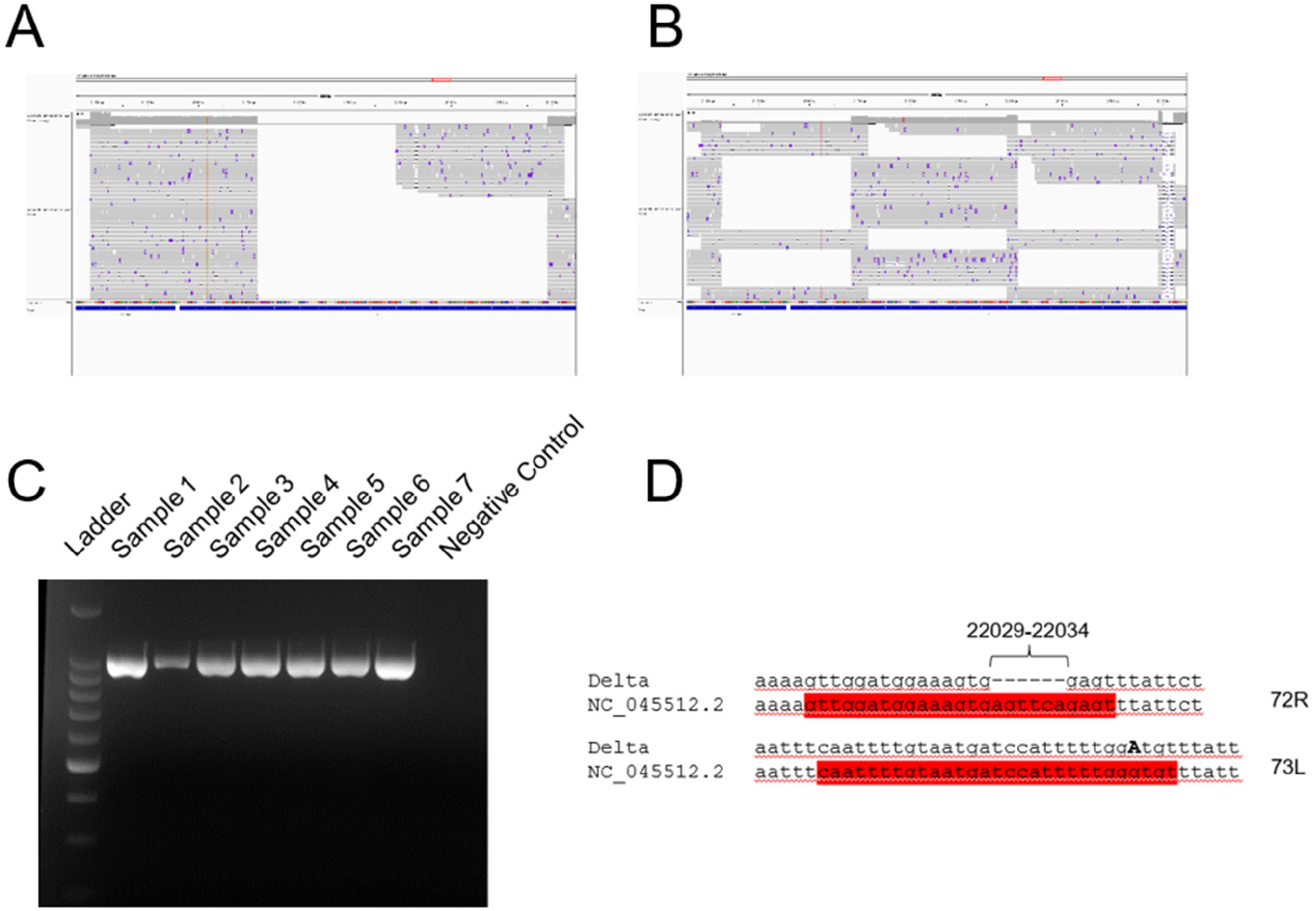
**A, B**) Coverage analysis of the ROI (21357-22346 nt) for representative Delta variant (A) and lambda variant (B) reads. Snapshots were taken directly from the visualization of the BAM files with IGV software. **C**) Agarose gel electrophoresis of PCR amplicons generated with primers 71L and 73R on Lambda (samples 1-2), Delta (samples 3-7), and negative (well 8) samples. Ladder: 1-kb ladder. **D**) Observed mutations at primer binding sites for primers 72R and 73L. The two primers are highlighted in red and the substitution G21987A is indicated in bold, capital letter.

### The USR can be observed genome sequences from many countries

We further investigated SARS-CoV-2 sequences originating from six countries (England, France, Germany, India, Switzerland, USA) to assess the presence of N-containing sequences in the ROI by looking at all sequences available in the GISAID database corresponding to Delta variants until 12 August 2021 (i.e. B.1.167.2, AY.1, AY.2, AY.3). For each sequence, we determined the proportion of Ns present in the ROI as compared to that in the rest of the sequence, by using PCR *in silico* with primers 71L and 73R. As we were interested in longer USRs that would originate from PCR mis-amplification than in small segments containing N bases, we filtered out all N-containing segments smaller than 200 nt and removed sequences with low coverage (overall percentage of N > 5%), in accordance with the criteria used by GISAID criteria. The mapping of large N-containing segments for each genome clearly indicated that a large portion of these segments were located in the ROI for sequences originating from all six countries considered (**Fig. 3A**), especially when retaining N-containing segments larger than 300-nt segments (**Fig. 3B**). Another striking observation was the large amounts of N-containing segments outside the ROI corresponding to other USR regions, particularly in countries contributing large amounts of sequencing data (USA, England). We also observed that the contribution of the USR in the S region to the total number of Ns in each genome sequence was disproportionally high (about 50% and higher), and started already from the early reported cases of Delta sequences in April-May 2021 across all countries considered here and continued to fluctuate markedly in all countries (**Fig. 4**). Altogether, our observations indicate that there is a clear coverage bias particularly marked in this ROI, and that the issue is generalized both geographically and across time.

**Figure 3.**
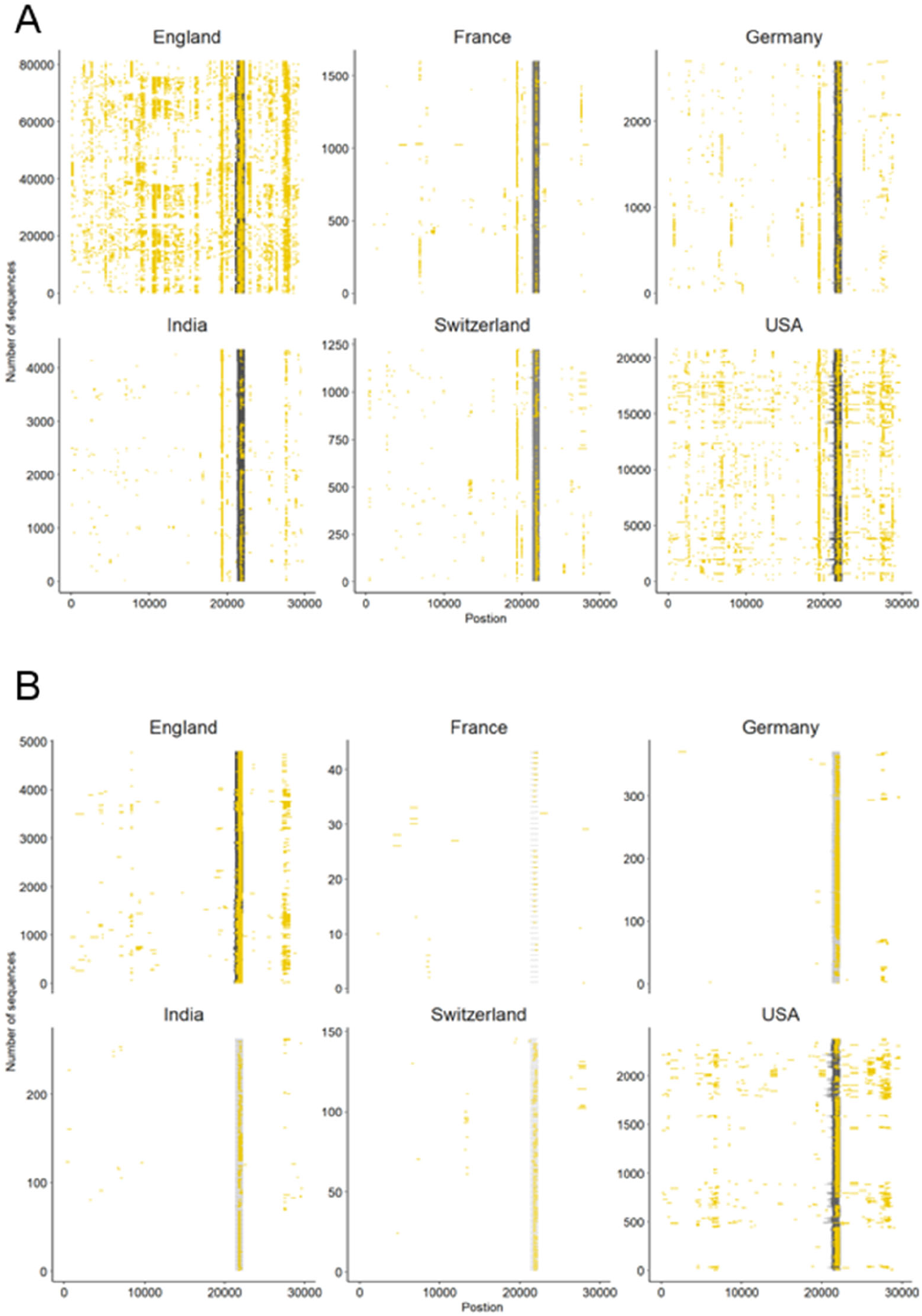
Occurrence of N-containing segments in SARS-CoV-2 genome sequences. Each N-containing segment is depicted in yellow and is positioned on the horizontal axis according to its position within the consensus sequence. The vertical axis represents the number of consensus sequences analysed among six countries representative of the global situation. Only sequences with overall less than 5% Ns and **A**) N-containing segments of lengths >200 nt, and **B**) >300 nt are depicted.

**Figure 4.**
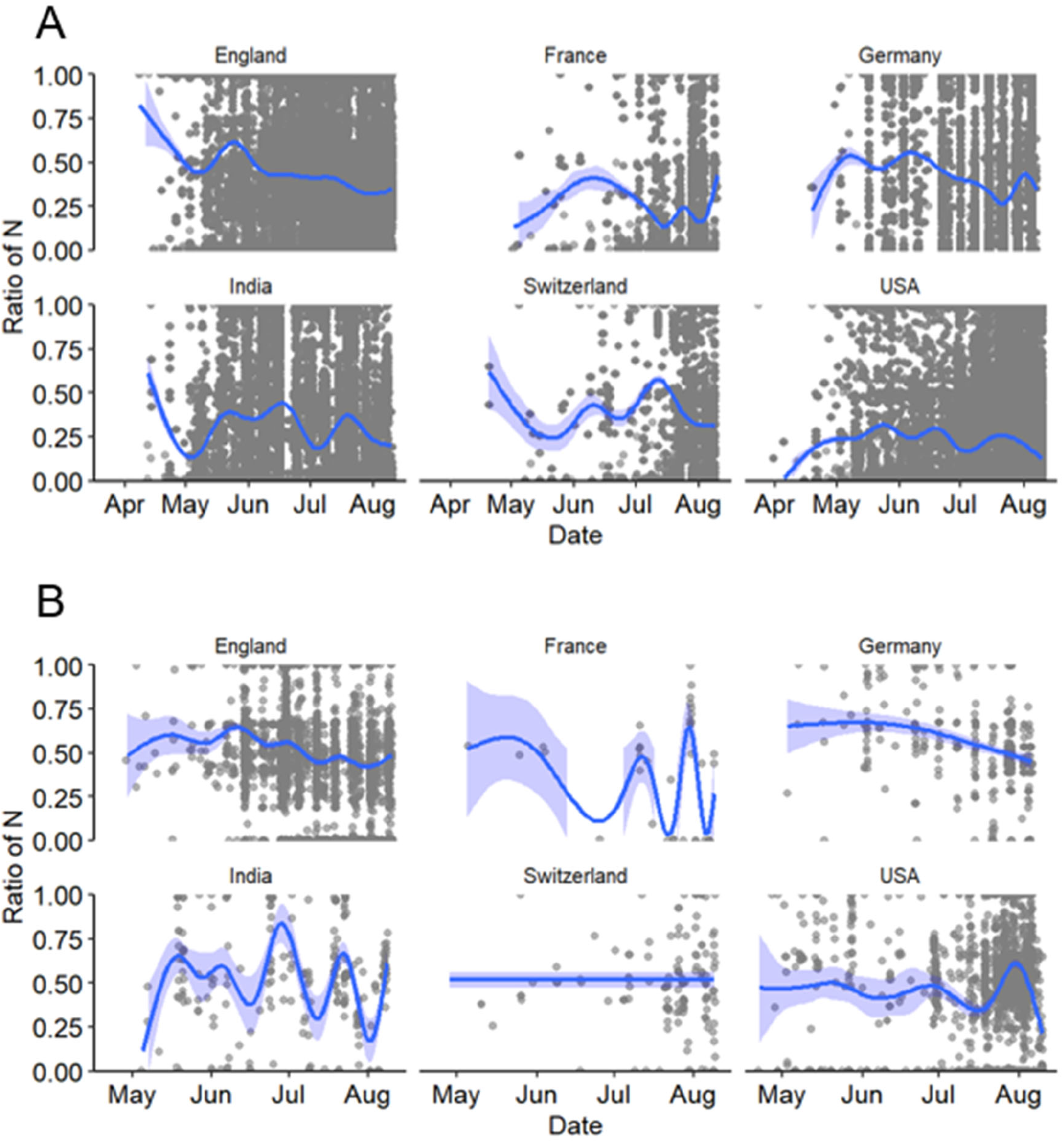
Ratio of number of Ns present in the ROI to the total number of Ns present in the entire genome of SARS-CoV-2 sequences for six countries in 2021. Considered here are **A**) all sequences with a total <5% of Ns and **B**) those with N-containing segments of length > 200 nt. The dark blue lines depict the best fitting lines of generalized additive models (GAM) with 95% confidence intervals (light blue areas).

### Design of Delta-specific primers

Because primer mismatch issues can lead to lineage miss-classification and to missing mutations of interest in the spike region, we modified the two primer sequences (72R and 73L) that display matching issues. For primer 72R, we created three alternative primers (72R_alt1, 72R_alt2 and 72R_alt3), and for 73L, another three primers (73L_alt1, 73L_alt2 and 73L_alt3) (**Fig.5A, Table 1)**. All new primers were successfully validated by PCR done in isolation from other primers (**Fig. 5 B**). Next, to confirm that the newly generated primers increased the coverage of the ROI, we added them to the original pool of Artic v3 primers. We chose four different samples comprising three different lineages (B.1.617.2, AY.4 and AY.12). Two different concentrations were used to test the new primers: 1) The primers were added in the same concentration as the other ARTIC primers (i.e. 0.5 μl of 100 mM primer solution), or as 2) twice the standard primer concentrations to favour their use during PCR amplification. First, we observed an increase in coverage for both B.1.617.2 samples (Sample 1 and Sample 2) when using the modified pool of primers **(Fig. 6)**. In Sample 3 (AY.4), we observed a clear enhancement of the coverage in the ROI, especially with the higher concentration of primers (**Fig. 6**). Unfortunately, in Sample 4 (AY.12) the region between nucleotide 21716 and 21989 displayed no read at all. These results indicate that the modifications of the primers may allow to recover complete coverage of Delta variants of lineages B.1.617.2 and AY.4 but not in AY.12, at least for the samples tested.

**Figure 5.**
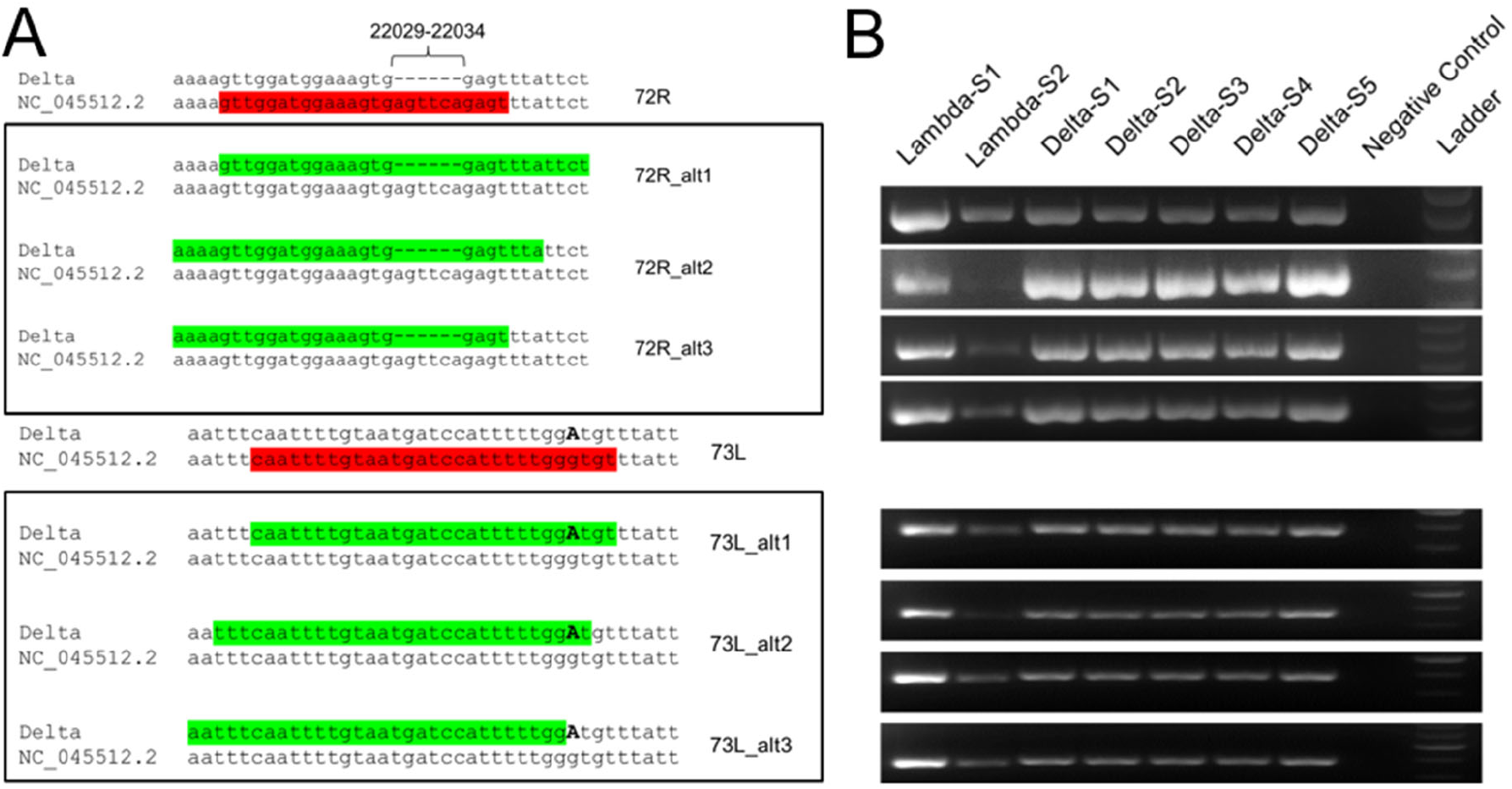
Sequence analysis of proposed primers. **A**) Alternative primers for 72R (72R_alt1, 72R_alt2, 72R_alt3) and for 73L (73R_alt1, 73R_alt2, 73R_alt3) primers were generated with Delta variant sequences as template, and aligned back to the NC_045512.2 reference sequence. The original primers are indicated in red font, and modified primers in green font, highlighting the presence of missing bases and substitution (G21987A). **B**) The alternative primers were validated by PCR under the same conditions as the ones used in the ARTIC v3 protocol.

**Figure 6.**
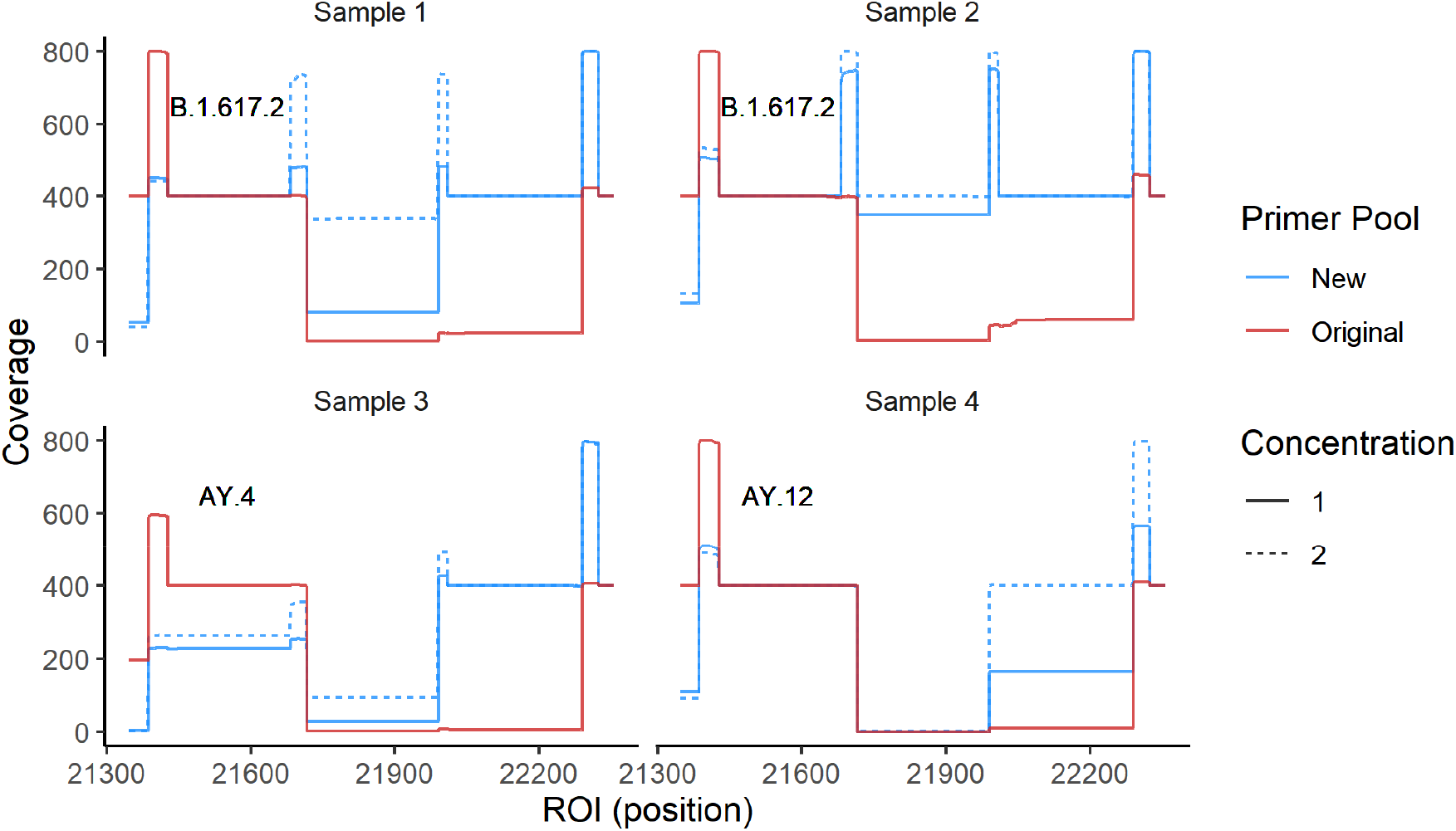
Coverage of the ROI using primer pools containing Delta-specific primers. We sequenced four samples of three different lineages using two pools of primers: 1) The original ARTIC v3 and 2) the ARTIC v3 pools where we added all newly designed primers in two concentrations, with concentration 1 corresponding to the standard primer concentration, and concentration 2 to twice that amount. The coverage using the new pool of primers (blue) and the original pool (red) are shown.

## Discussion

In the context of routine SARS-CoV-2 genomic sequencing, we observed the presence of a large under-sequenced region in the S gene of the viral genome. We identified that the issue originated from primer mismatch for ARTIC v3 primers 72R and 73L to the sequences of the now predominant Delta variants. The mismatch was caused by deletion and mutational events. We demonstrated that the presence of this USR in the ROI is a technical problem, which started early with the first appearance of Delta variants worldwide, and still continues to appear to large extent in the reported genome sequences globally. Finally, we provided a series of alternative primers for primers 72R and 73L, particularly addressing users of the ARTIC v3 protocol. These primers were validated and could be added to the large pools of the ARTIC v3 primers to ensure correct amplification and sequencing of the ROI. There is for sure an inherent difficulty to establish new, universal primer sequences given that the large amount of low-covered sequences may hide some unknown sequence diversity in the ROI.

To our knowledge the systematic presence of this USR has not been reported elsewhere. Although we could identify and provide a solution for this ROI, our analysis of data at the global scale indicated that many USR may also be present in other regions of the genome of SARS-CoV-2. Therefore, it is of major importance to regularly control for the presence of USRs and determine how geographically and temporally consistent they become. Although it may be easier to track the wet laboratory protocols used for SARS-CoV-2 genomic sequencing for countries reporting smaller data volumes, it may be less so for countries providing large amounts of data from many sequencing facilities. Currently little information is available in the sequence metadata about some of the key procedures involved in the generation of SARS-CoV-2 genome sequences, such as reverse transcription conditions, choice of primer sets, PCR amplification conditions, etc., and more focus is rather given to sequencing technologies. We encourage genome data submitters to provide wet laboratory information in the future to allow for higher methodological comparability and for identifying protocols that would need improvement.

## Data Availability

The study is based on publicly available data at GISAID (https://www.epicov.org/).

